# Memory Self-Awareness Phenotypes are Associated with Alzheimer’s Disease Biomarker Burden and Clinical Progression in Amnestic Mild Cognitive Impairment

**DOI:** 10.64898/2026.11.03.26367187

**Authors:** Nicholas Enbar-Salo, Phebe Palmer, Catherine Munro, Mark Dubbelman, Anna Aaronson, Rachel Nosheny, Jennifer Gatchel, Gad A. Marshall, Patrizia Vannini, the Alzheimer’s Disease Neuroimaging Initiative

## Abstract

**Background:** Altered memory self-awareness is common in mild cognitive impairment (MCI), but its relationship to Alzheimer’s disease (AD) pathology and progression remains unclear.

**Methods:** Using 590 amnestic MCI ADNI participants, memory self-awareness was derived from participant–informant discrepancies on the memory subdomain of the Everyday Cognition questionnaire. Heightened awareness and unawareness subscores were calculated, and k-means clustering identified four phenotypes. Associations with cerebrospinal fluid (CSF) and positron emission tomography (PET) were examined, and clinical progression was assessed using Cox models.

**Results:** Aware and Unaware groups showed lower CSF amyloid-β, and higher CSF phosphorylated tau and PET amyloid. The Unaware phenotype was associated with increased progression risk (hazard ratio 2.41, 95% CI 1.48–3.92), whereas the Worried Well phenotype showed reduced risk (hazard ratio 0.58, 95% CI 0.34–0.99).

**Conclusions:** Multidimensional assessment of memory self-awareness identifies biologically and prognostically distinct subgroups within MCI and may improve risk stratification in prodromal AD.

## BACKGROUND

Alzheimer’s disease (AD) is the leading cause of cognitive impairment among older adults and is frequently accompanied by altered awareness of cognitive decline. Self-awareness of cognitive function is commonly assessed by examining discrepancies between participant- and study partner–reported cognitive complaints [1, 2]. Heightened awareness, self-perceived decline not corroborated by an informant [3], has often been observed in preclinical stages, whereas unawareness (anosognosia), defined as greater informant-reported than self-reported decline, is characteristic of AD dementia [4, 5]. In mild cognitive impairment (MCI), awareness is heterogeneous, with individuals ranging from intact insight to marked unawareness [4–6]. Estimates of anosognosia in MCI vary widely[1, 7–15], underscoring uncertainty regarding its clinical and biological significance.

This heterogeneity has important implications. Reliance on participant-reported subjective cognitive decline alone may be misleading when awareness is impaired, highlighting the importance of incorporating study partner reports in early disease stages [5]. Because episodic memory impairment is the defining clinical feature of amnestic MCI, and one of the earliest cognitive manifestations of AD, memory self-awareness represents a clinically and biologically relevant construct for investigating heterogeneity in disease progression. As efforts intensify to identify individuals at highest risk of progression, particularly in the era of disease-modifying therapies, the pathological and prognostic relevance of altered self-awareness in MCI has become increasingly important. Despite these efforts, methods to effectively stratify individuals within the MCI stage remain limited – a gap that may be addressed by leveraging heterogeneity of memory self-awareness.

Traditional discrepancy scores average differences across questionnaire items, potentially obscuring distinct patterns of heightened awareness and unawareness within individuals. A recently proposed approach separates these dimensions by independently quantifying unawareness and heightened awareness based on item-level discrepancies [16]. For each questionnaire item, discrepancies are calculated between participant and study partner ratings; items on which the study partner reports greater decline contribute to an unawareness subscore, whereas items on which the participant reports greater decline contribute to a heightened awareness subscore. By isolating these directional discrepancies rather than averaging them into a single summary score, this approach yields a two-dimensional construct that distinguishes loss of awareness from increased concern. In cognitively normal older adults from the Alzheimer’s Disease Neuroimaging Initiative (ADNI), only the unawareness subscore, rather than the traditional discrepancy score or the heightened awareness subscore, was associated with future clinical progression [16]. These findings are consistent with prior studies demonstrating that reduced awareness, but not heightened concern, is linked to greater clinical decline [17, 18], suggesting that directional measures of self-awareness may capture clinically meaningful risk. Importantly, these studies have primarily focused on memory self-awareness, reflecting the central role of episodic memory impairment across the AD continuum and providing the rationale for the present study’s focus on memory self-awareness. However, whether these constructs reflect underlying AD pathology or predict progression once cognitive impairment is present remains unknown. Moreover, traditional discrepancy measures do not account for the overall magnitude of participant and informant complaints. For example, a discrepancy score of zero may reflect either low complaints from both individuals or high but concordant complaints (i.e. when the participant and informant report similar levels of decline), potentially obscuring clinically meaningful differences in risk [19, 20].

Data-driven approaches may offer greater sensitivity for identifying distinct self-awareness profiles. Unsupervised clustering methods allow integration of participant-reported complaints, study partner– reported complaints, and independent measures of unawareness and heightened awareness to derive clinically interpretable phenotypes [21, 22].

In this study, we sought to characterize memory self-awareness phenotypes in individuals with amnestic MCI using a clustering approach and to examine their associations with cerebrospinal fluid (CSF) biomarkers of amyloid and tau and with longitudinal progression to AD dementia. Because amnestic MCI is defined by impairment in episodic memory, the cognitive domain most closely linked to the earliest stages of AD pathology, and previous studies of altered self-awareness in Alzheimer’s disease have primarily focused on memory function, we examined memory self-awareness as our primary construct of interest. We hypothesized that phenotypes characterized by greater study partner–reported decline would demonstrate higher AD biomarker burden and increased risk of clinical progression [18].

## METHODS

### Participants

All participants (N = 590) used in the preparation of this article were MCI at baseline and were obtained from the ADNI database (adni.loni.usc.edu). ADNI was launched in 2003 as a public-private partnership, led by Principal Investigator Michael W. Weiner, MD, and includes the extensions ADNI-GO (launched 2009), ADNI-2 (2011), and ADNI-3 (2016). The original goal of ADNI was to test whether serial magnetic resonance imaging (MRI), positron emission tomography (PET), other biological markers, and clinical and neuropsychological assessments can be combined to measure the progression of MCI and AD dementia. The current goals include validating biomarkers for clinical trials, improving the generalizability of ADNI data by increasing diversity in the participant cohort, and providing data concerning the diagnosis and progression of AD to the scientific community. For up-to-date information, see adni.loni.usc.edu. Exclusion criteria included a history of alcoholism, drug abuse, and head trauma, as well as serious medical or psychiatric conditions. Cholinesterase inhibitors and memantine were only allowed in participants if they were stable for 3 months before screening. Moreover, antidepressants were allowed if participants were not significantly depressed at the time of screening and did not have a history of major depressive disorder within the past year. Institutional review board approvals and informed consent were obtained prior to all procedures. This manuscript conforms to the Strengthening the Reporting of Observational Studies in Epidemiology ([STROBE](http://equator-network.org/reporting-guidelines/strobe/)) reporting guidelines for cohort studies.

The selection criteria for this study required both a participant and study partner response on the memory component of the ECog questionnaire from the same time point, as well as a clinical examination and a lumbar puncture to collect CSF occurring within a year of ECog completion. Their clinical diagnosis was used as a diagnostic criterion for amnestic MCI, determined by a Clinical Dementia Rating (CDR) Scale global score of 0.5, with a Memory Box score of at least 0.5, a Mini-Mental State Exam (MMSE) score between 24 and 30 inclusive, and performance below education-adjusted cutoffs on the Wechsler Logical Memory II subscale. For the survival analyses, a diagnosis of AD dementia was assigned to participants with a CDR global score of ≥ 0.5, an MMSE score between 20 and 24, performance below education-adjusted cutoffs on the Wechsler Logical Memory II subscale, and a probable AD label according to NINCDS/ADRDA criteria. The participants included individuals sampled from the ADNI-GO, ADNI-2, and ADNI-3 phases, spanning from 2010 to 2022. ADNI-1 was not included in the sample since the ECog questionnaire was administered starting in the ADNI-GO phase in 2010. Any individuals with missing baseline ECog questionnaire and CSF biomarker data were removed from the analysis, and any longitudinal observations without clinical diagnosis data were removed. For the survival analysis, we created a subset that included participants with at least two follow-up clinical diagnosis time points (N = 492). For more information on ADNI, see the ADNI database (adni.loni.usc.edu).

### Self-awareness measure

Memory self-awareness was measured using the memory subsection of the ECog Questionnaire. The ECog is comprised of 39 items, 8 of which specifically focus on memory [23, 24]. The memory subsection primarily evaluates episodic memory, with participants and their study partners independently assessing the participant’s ability to accomplish everyday memory tasks compared to 10 years earlier, such as remembering a few shopping items without a list, or if there has been any change in repeating stories and/or questions. Responses were recorded on a Likert scale, where 1 indicated improvement or no change, 2 reflected occasional or questionable decline, 3 denoted consistent mild decline, and 4 signified consistent significant decline. An “I do not know” option was also available and treated as missing data when selected.

A two-step approach was used to assess self-awareness. First, an unawareness sub-score and a heightened awareness sub-score for the ECog Questionnaire, originally published in [16] were calculated for each participant. The unawareness sub-score was calculated by subtracting the study partner’s ratings from the participant’s ratings for each item, setting all positive differences to zero, and averaging the results (including the zeros derived from the positive differences). Similarly, the heightened awareness sub-score was calculated by capping all negative differences at zero and taking the mean, again including the zeroes derived from negative differences in the mean computation. Second, k-means clustering (with Euclidian distance) using the kmeans package was used to categorize participants into four distinct clusters, with 25 random initializations to ensure stability and the seed set to 42. K-means was chosen due to the continuous nature of the ECog questionnaire. Additionally, k-means clustering is uniquely fast, scalable, and intuitive by assigning all individuals into discrete clusters (unlike probabilistic methods such as Latent Profile Analysis). These factors make k-means easy to integrate into clinical settings for risk stratification. Prior to clustering, all four features were z-scored, and nine outliers based on the four awareness measures were discovered using Mahalanobis distance and excluded from clustering. Mahalanobis distance was used due to its ability to check for multivariate outliers. Assumptions of sphericity for k-means clustering were checked visually using principal component analysis. The choice of using four clusters was determined with the elbow method and the Davies-Bouldin Index (DBI; see **eFigures 1** and **2**), along with prior literature, which has used both participant and study partner scores from the ECog questionnaire at a cut off of 2.5 for both measures to classify individuals into the following awareness quadrant profiles: 1) Low Complaints, 2) Worried Well, 3) Aware, and 4) Unaware [25]. Four measures were used as clustering features: the unawareness sub-score, the heightened awareness sub-score, the mean study partner complaints, and the mean participant complaints. We evaluated the quality of clustering using the Davies-Bouldin Index **(eFigure 2)** and silhouette scores. These metrics were chosen as they are shown to be more informative and reliable compared to alternatives (e.g. Shannon entropy, Dunn index) in biomedical contexts [26]. The Davies-Bouldin Index (DBI) is a measure of the average ratio of within-cluster scatter and between-cluster separation. Scores on the DBI range from zero to positive infinity, with lower scores indicating more compact, well-separated clusters [27]. Meanwhile, the silhouette score measures how similar each point is in Euclidean space to other clusters. Silhouette scores range from -1 to +1, with scores closer to +1 indicating better clustering quality [28]. Silhouette scores were computed for each cluster to capture inter-cluster variability in cluster quality that can be masked when looking at a global silhouette score. Due to the multicollinearity of our features, we also used these four measures in principal component analysis (PCA) to create clusters with orthogonal features. This PCA found that two components accounted for 90.1% of the variance between our features.

### CSF Measurements of AD Core Biomarkers

Lumbar punctures were performed as part of the ADNI protocols and CSF was collected, centrifuged, subpackaged, and stocked in polypropylene tubes in a refrigerator at -80 °C. ADNI used the Roche Elecsys immunoassay to assess AD biomarkers, including total tau (t-tau), phosphorylated tau (p-tau) (181P) and Aβ (1-42). A single lot of reagents for each of the three measured biomarkers was collected (provided in UPENNBIOMK9.csv file available at http://adni.loni.usc.edu). CSF was collected both at baseline and follow-up visits. The following procedure only used p-tau and Aβ1-42 as measures for tau and amyloid, respectively. CSF was chosen over other measures due to PET, blood-based, and plasma measures only becoming standard in ADNI, which would have limited statistical power and follow-up duration for many participants. However, supplemental analyses were performed in a subsample (N = 114) with PET inferior temporal tau standard uptake value ratios (SUVR), entorhinal tau SUVR, and amyloid centiloids.

### Statistical Analyses

All analyses were conducted in R version 4.4.1 (R Project for Statistical Computing). The threshold for statistical significance was *p* < .05.

Given the non-linear distributions of both CSF biomarkers, we utilized Kruskal–Wallis tests to assess cross-sectional differences in CSF biomarkers among the four awareness phenotypes. Significant omnibus effects were followed by post hoc pairwise Wilcoxon rank sum tests to identify specific group differences. To control for multiple comparisons, p-values were adjusted using the Benjamini–Hochberg procedure for false discovery rate correction where applicable. To account for age, sex, and education, we conducted confirmatory Rank-Based analyses of covariance (ANCOVAs) using the Rfit packages, using CSF measures in two separate tests as predictors and with cluster, age, sex, and education as covariates.We used post hoc Wilcoxon rank sum tests on the residuals of the covariates to look for specific group differences. Supplemental analyses also performed these tests with PET tau (inferior temporal and entorhinal) and amyloid centiloids as predictors.

Survival analyses were run using Cox regression models using the survival package to examine the longitudinal associations of each awareness phenotype at baseline and time to progression (measured in years from baseline). Outcome variables were progression status and (1) time to progression, for progressors, and (2) total time observed, for stable (i.e., censored) individuals. Progression from MCI to AD dementia was determined using progression by clinical diagnosis. The model was run with phenotype membership of each participant as the primary predictor, and the model adjusted for baseline age, sex, and education. The Cox proportional-hazard model was used to find correlation among the factors, and hazard ratio (HR) and 95% confidence interval (CI) were calculated. To assess the fit of these progression analyses, Uno’s C-index, time-dependent AUC values at times 1, 2, and 4 (corresponding to each quartile of the proportion of progression events), and the inverse probability of censoring weighted (IPCW) Brier scores were computed. To ensure that any clinical or biological differences in phenotypes were not due to underlying clinical impact severity, supplemental sensitivity analyses were also controlled for MMSE.

## RESULTS

### Clustering and Demographics

Overall, four phenotypes were created (**Figure 1**). Cluster one, the “Low Complaints” phenotype, had low scores in all measures. Cluster two, the “Unaware” phenotype, was characterized by a high unawareness sub-score and high complaints from the study partner, along with low scores in heightened awareness and low complaints from the participant. Cluster three, the “Aware” phenotype, had low heightened awareness and unawareness scores, and high complaint magnitude from both the participant and study partner. Finally, cluster four, the “Worried Well” phenotype, had high heightened awareness, low unawareness, high participant complaints, and low study partner complaints. See **Figure 1** for the direction and magnitude of all four awareness measures used to construct the phenotypes. With a DBI score of 0.936, the clustering was a good statistical fit for the data (**eFigure 2**), and all assumptions for k-means clustering were met. The per-cluster mean silhouette scores ranged from 0.293 for the Aware phenotype and 0.422 for the Unaware phenotype, indicating modest cluster separation.

**Figure 1.**
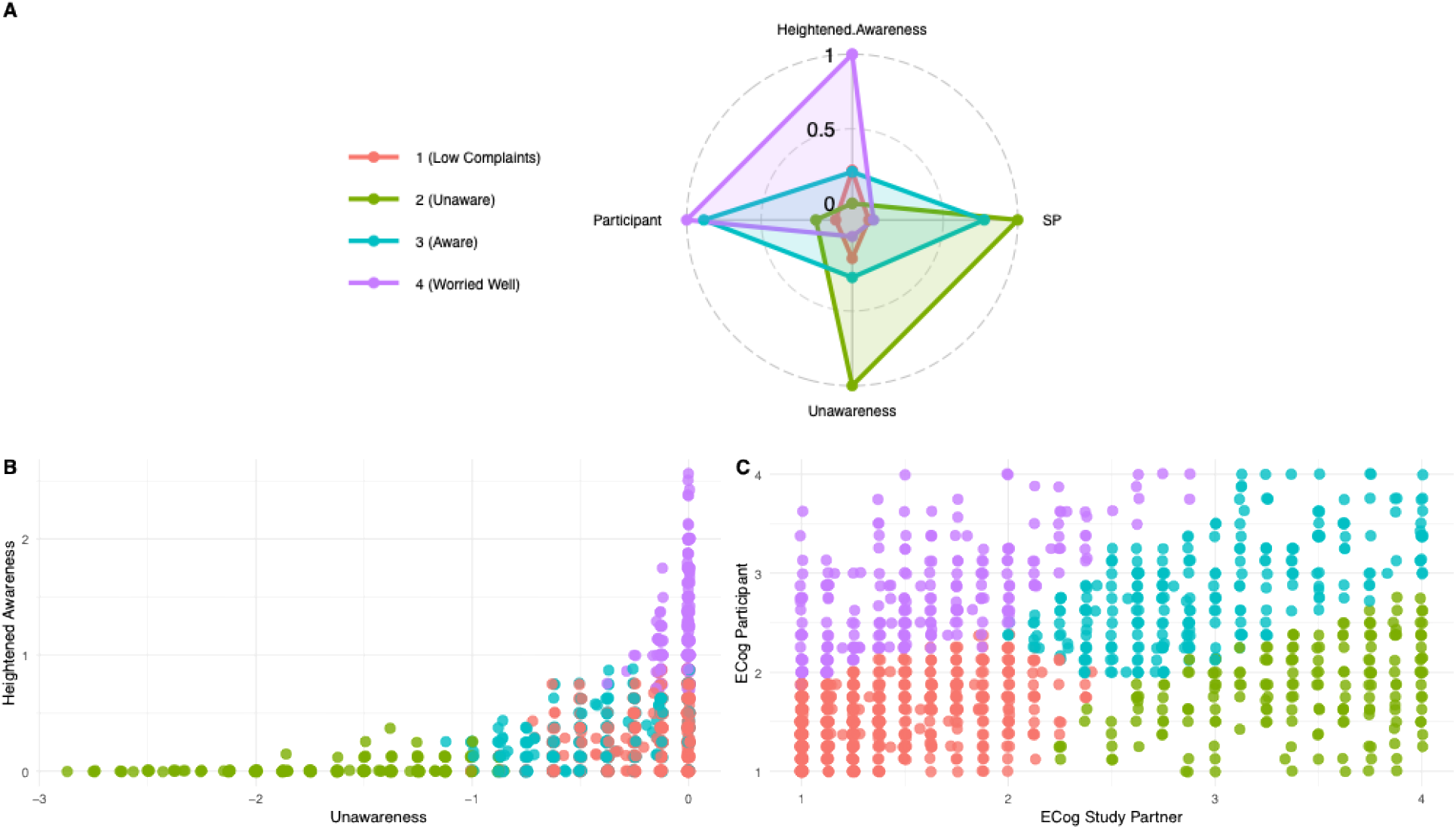
Mean and Distribution of the Clustering Parameters by Cluster. Radar map and scatter plots for (A) the means of the measures used to cluster, (B) the distribution of clusters by the unawareness and heightened awareness subscores, and (C) the distribution by study partner and participant complaints. For the radar map, SP is an abbreviation for study partner, and all means are on a 0-1 scale, where 1 represents the maximum mean for each measure.

The sample was comprised of 260 females (43%) and 339 males (57%), with a mean (SD) age of 72.0 (7.6) and years of education of 16.22 (2.6). The sample consisted primarily of White individuals (93%) and included 19 Hispanic or Latino individuals (3.2%). The mean (SD) for the MMSE was 28.0 (2.0), and depression (as assessed with the Geriatric Depression Scale) was low (mean (SD) of 1.8 (1.5)). The rate of progression to AD dementia significantly differed by cluster (p < 0.001), with the highest proportion in the Unaware phenotype (29%) and the lowest in the Worried Well phenotype (5.2%). Years to progression also differed significantly across phenotypes (p = 0.04), with the Unaware group having the least years observed, and the Worried Well phenotype having the most (**Table 1**). Among the 114 participants in the PET subsample, 34 were classified as Low Complaints, 15 as Unaware, 36 as Aware, and 29 as Worried Well, with demographic and clinical means in this subsample closely resembling those of the entire sample.

**Table 1.** Demographics and clinical characteristics for the sample.

| **Characteristic** | **All<br>N = 590<sup>a</sup>** | **Low Complaints<br>N = 184<sup>a</sup>** | **Unaware<br>N = 82<sup>a</sup>** | **Aware<br>N = 171<sup>a</sup>** | **Worried Well<br>N = 153<sup>a</sup>** | **p-value<sup>b</sup>** |
|---|---|---|---|---|---|---|
| Age | 72.0 (7.5) | 71.6 (7.4) | 73.6 (7.3) | 72.4 (7.4) | 71.1 (7.8) | 0.04* |
| Gender (F) (No. %) | 257 (44) | 80 (43) | 32 (39) | 73 (43) | 72 (47) | 0.68 |
| Years of Education | 16.2 (2.6) | 16.3 (2.4) | 16.1 (2.6) | 16.0 (2.8) | 16.3 (2.7) | 0.80 |
| Ethnicity (Hispanic or Latino) (No. %) | 19 (3.2) | 8 (4.3) | 4 (4.9) | 6 (3.5) | 1 (0.7) | 0.12 |
| Race (White) (No. %) | 549 (93) | 172 (93) | 76 (93) | 163 (95) | 138 (90) | 0.14 |
| MMSE | 28.0 (2.0) | 28.5 (1.6) | 27.3 (2.5) | 27.7 (2.1) | 28.0 (1.8) | <0.001*** |
| GDS | 1.8 (1.5) | 1.3 (1.5) | 1.5 (1.4) | 2.2 (1.5) | 2.1 (1.5) | <0.001*** |
| ECog Participant Score | 2.3 (0.7) | 1.7 (0.3) | 1.8 (0.4) | 2.7 (0.5) | 2.9 (0.5) | <0.001*** |
| ECog Study Partner Score | 2.2 (0.8) | 1.6 (0.4) | 3.2 (0.5) | 2.9 (0.5) | 1.7 (0.4) | <0.001*** |
| Unawareness Sub-Score | -0.4 (0.5) | -0.2 (0.2) | -1.4 (0.4) | -0.4 (0.3) | 0.0 (0.1) | <0.001*** |
| Heightened Awareness Sub-Score | 0.5 (0.5) | 0.3 (0.2) | 0.0 (0.1) | 0.3 (0.2) | 1.2 (0.4) | <0.001*** |
| Progressed to AD (No. %) | 81 (14) | 15 (8.2) | 24 (29) | 34 (20) | 8 (5.2) | <0.001*** |
| Years to Progression | 2.8 (2.2) | 3.0 (1.9) | 2.0 (1.2) | 2.7 (2.1) | 5.5 (3.2) | 0.04* |

<sub>a</sub>Mean (SD); n (%)
<sub>b</sub>Kruskal-Wallis rank sum test; Pearson's Chi-squared test; Fisher's exact test

### CSF and PET Pathology Across Memory Self-Awareness Phenotypes

To investigate the association between each memory self-awareness phenotype and CSF biomarkers of amyloid and tau, Kruskal-Wallis tests were conducted. Both CSF Aβ1-42 (H(3) = 13.59; p = 0.004) and p-tau (H(3) = 13.09; p = 0.004), as well as PET amyloid centiloids (H(3) = 19.26, p < 0.001), were significant independent predictors among the clusters (**Table 2**). However, no such significance was found for either PET entorhinal or inferior temporal tau (Table 2**; eFigure3b** and **eFigure3c**). For between-group differences for CSF Aβ1-42, post hoc Wilcoxon Rank-Sum tests revealed that both the Unaware and Aware phenotypes had significantly lower Aβ1-42values compared to the Worried Well (p = 0.037 for both phenotypes) and Low Complaints phenotypes (p = 0.020 for both phenotypes, **eFigure 4a**). Meanwhile, for p-tau (**eFigure 4b**), the Unaware phenotypes had significantly greater p-tau values than the Worried Well (p = 0.030) and Low Complaints phenotypes (p = 0.007), but the Aware phenotype was only significantly greater than the Low Complaints phenotype (p = 0.030). For both CSF measures, non-significant p-values were found between the Unaware and Aware phenotypes, or the Worried Well and Low Complaints phenotypes. For PET amyloid centiloids (**eFigure 3a**), the Aware phenotype was found to have significantly greater centiloid values compared to the Low Complaints (p = 0.013) and the Worried Well (p < 0.001) phenotypes, while the Unaware phenotype was only significantly greater than the Low Complaints phenotype (p = 0.012). Confirmatory Rank-Based ANCOVA results, correcting for sex, age, and education, found similar findings to the Kruskal-Wallis tests. Similar findings were observed after controlling for cognitive performance and when PCA-derived clusters were used.

**Table 2.**
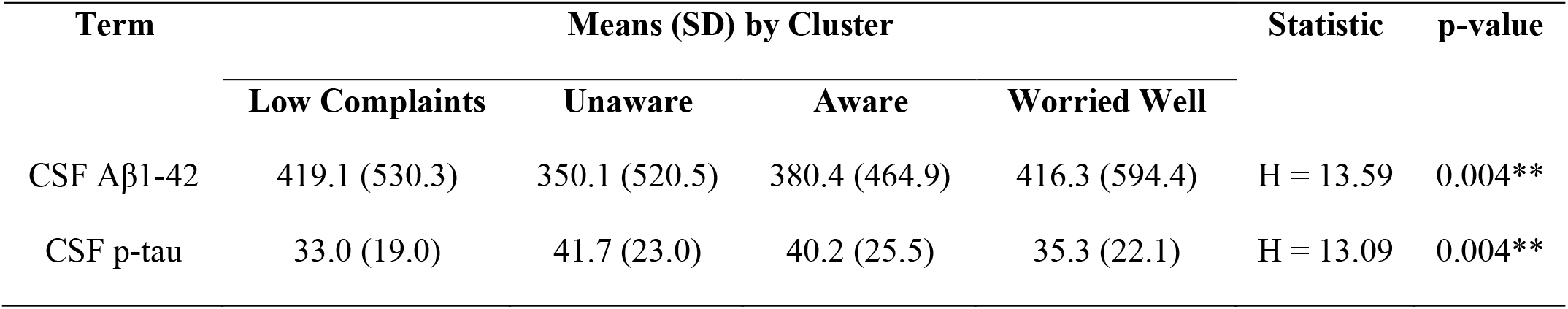
Means (SD) and Association of awareness phenotype with CSF Aβ_1-42_ and p-tau for MCI individuals.

| **Term** | **Means (SD) by Cluster** |  |  |  | **Statistic** | **p-value** |
|---|---|---|---|---|---|---|
|  | **Low Complaints** | **Unaware** | **Aware** | **Worried Well** |  |  |
| CSF Aβ1-42 | 419.1 (530.3) | 350.1 (520.5) | 380.4 (464.9) | 416.3 (594.4) | H = 13.59 | 0.004** |
| CSF p-tau | 33.0 (19.0) | 41.7 (23.0) | 40.2 (25.5) | 35.3 (22.1) | H = 13.09 | 0.004** |

### Survival Analysis Across Clusters

To analyze the effect of each memory self-awareness phenotype on clinical progression in individuals who had an MCI diagnosis at baseline, we performed Cox regression analyses, correcting for age, gender, and education. All clusters were found to be significant when compared to the Low Complaints cluster as a reference (**Table 3**; **Figure 2**). The Worried Well phenotype, compared to the Low Complaints phenotype, was associated with a 42% decrease in progression hazard (HR ratio = 0.58, 95% CI: 0.34, 0.99; p = 0.044). Meanwhile, the Aware and Unaware phenotypes were associated with increased progression, with the Aware phenotype being associated with a 64% increase in progression hazard (HR ratio = 1.64, 95% CI: 1.06, 2.52; p = 0.026), and the Unaware phenotype being associated with a 141% increase in progression hazard as compared to the Low Complaints phenotype (HR ratio = 2.41, 95% CI: 1.48, 3.92; p < 0.001). However, when controlling for cognitive performance, the Aware phenotype was no longer significantly associated with an increase in progression risk (HR ratio = 1.48, 95% CI: 0.95, 2.31; p = 0.083). All other associations remained when controlling for MMSE. Furthermore, similar results were found when performing these analyses on PCA-derived clusters.

**Figure 2.**
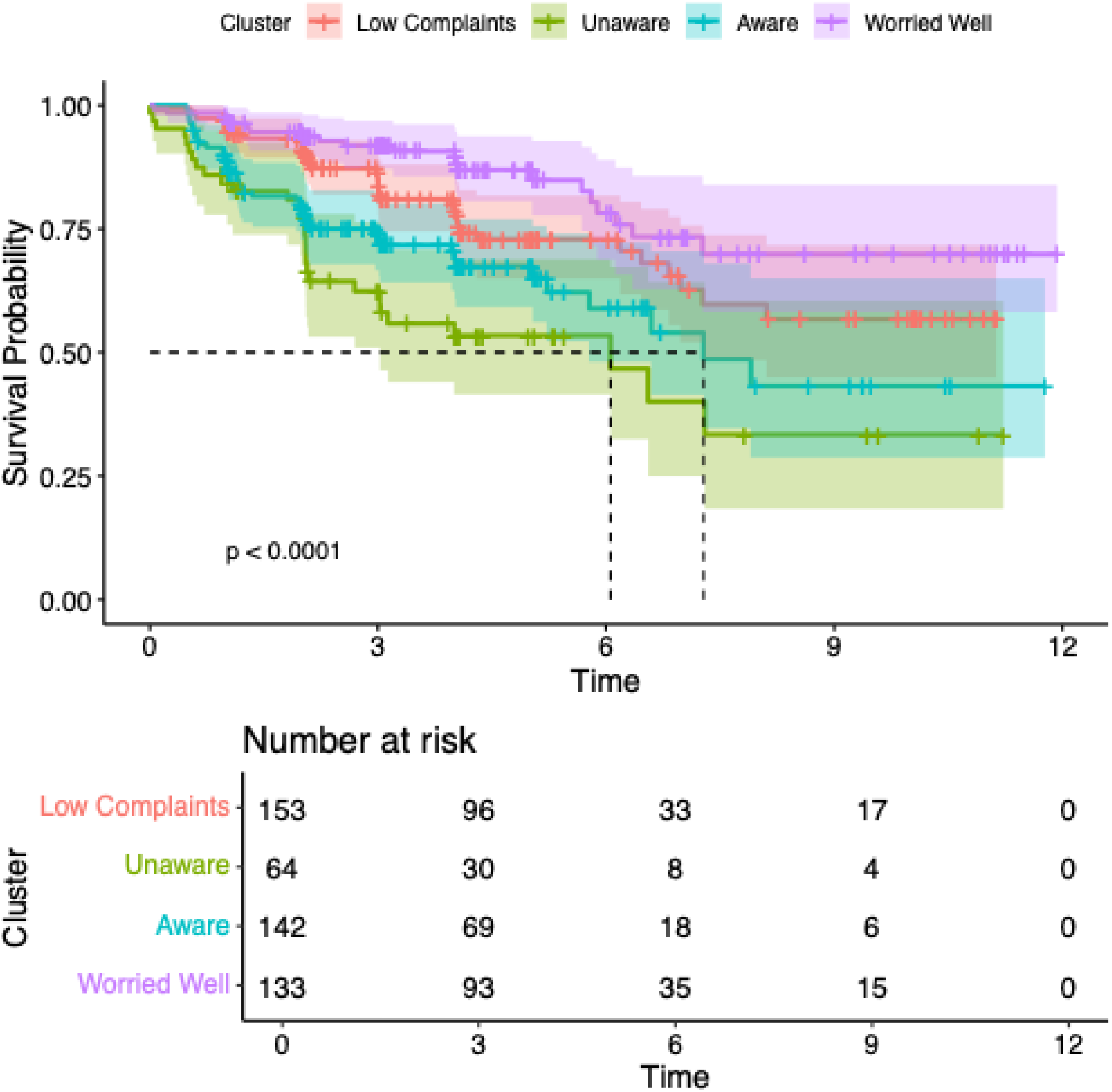
Kaplan-Meier Curves for Each Awareness Cluster Kaplan-Meier curve (above) and risk table by cluster (below). The Unaware phenotype is in green, and progressed the fastest of all phenotypes, followed by the Aware phenotype (in blue), the Low Complaints phenotype (in red), and the Worried Well phenotype (in purple).

**Table 3.** Survival Analysis Results for MCI individuals, with the Low Complaints cluster as the reference.

| **Variable** | ***b*** | **HR (95% CI)** | ***z*** | ***P* value** |
|---|---|---|---|---|
| Age | -0.01 | 0.99 (0.97 to 1.01) | -0.72 | 0.47 |
| Male Sex | -0.27 | 0.76 (0.53 to 1.08) | -1.53 | 0.13 |
| Years of Education | 0.03 | 1.03 (0.97 to 1.10) | 0.95 | 0.34 |
| Worried Well Cluster | -0.55 | 0.58 (0.34 to 0.99) | -2.01 | 0.044* |
| Aware Cluster | 0.49 | 1.64 (1.06 to 2.52) | 2.22 | 0.026* |
| Unaware Cluster | 0.88 | 2.41 (1.48 to 3.92) | 3.55 | <0.001*** |

All three groups met the proportional hazard assumptions. Uno’s C-index was computed to be 0.621 (95% CI: 0.565, 0.667), suggesting moderate discrimination. AUC’s were computed to be 0.769 at year 1, 0.753 at year 2, and 0.758 at year 4, and IPCW Brier score for our model was lower than that of the null model at all three time points (0.058 vs. 0.065 at 1 year, 0.108 vs. 0.124 at 2 years, and 0.150 vs. 0.179 at 4 years).

## DISCUSSION

In this cohort of individuals with MCI, distinct memory awareness phenotypes were associated with differential Alzheimer’s disease biomarker burden and markedly different risks of progression to dementia. Phenotypes characterized by greater study partner–reported decline, particularly the Unaware phenotype, demonstrated higher phosphorylated tau, lower amyloid-β concentrations, and substantially increased progression risk. In contrast, the Worried Well phenotype, marked by heightened self-reported concern without informant corroboration, was associated with lower risk of progression. These findings suggest that structured assessment of memory self-awareness using patient–informant discrepancies may provide clinically meaningful prognostic information during the prodromal stages of AD. Importantly, these findings should be interpreted within the context of memory awareness in amnestic MCI rather than as evidence that awareness operates uniformly across all cognitive domains.

Our results extend prior work demonstrating that unawareness predicts clinical decline in cognitively normal individuals [17, 18] by showing that directional measures of awareness remain informative once cognitive impairment is present. Notably, although the Unaware and Aware phenotypes did not significantly differ in cross-sectional biomarker burden, the Unaware phenotype conferred a substantially greater hazard of progression. This dissociation suggests that loss of awareness may capture aspects of disease vulnerability not fully reflected by baseline pathology alone. Importantly, informant reports alone would not distinguish between these two groups; rather, the interaction between self- and informant-reported decline appears to convey additional prognostic value above and beyond only informant report, which would be unable to separate the Unaware and Aware phenotypes.

Interestingly, age as a covariate was not found to have an association with risk progression. While this goes against most literature suggesting older age as a risk factor, we believe that, given the Unaware phenotype had the oldest mean age as well as the highest risk of progression, the risk that would have come with age might have been already captured via our phenotypes. Additionally, there is some literature using age as a covariate to find predictors of risk progression in the MCI stage that has found similar null findings for age [29, 30].

Additionally, our findings support a growing body of research demonstrating an association between loss of memory self-awareness and increased AD biomarker burden [3, 18, 31–39], reinforcing the biological relevance of unawareness across the disease continuum. The present study focused on memory self-awareness because participants met criteria for amnestic MCI, a syndrome defined by impairment in episodic memory, which represents the earliest and most prominent cognitive manifestation of AD. Consequently, memory self-awareness was selected as the primary construct of interest rather than multidomain awareness. Whether the observed prognostic associations reflect a particular relevance of memory awareness in amnestic MCI or broader differences across cognitive domains remains an important question for future investigation and will require direct comparisons of domain-specific awareness measures.

The dissociation between cross-sectional biomarker burden and longitudinal progression risk in the Unaware phenotype raises important mechanistic considerations. Self-awareness of cognitive function depends on distributed neural networks involving medial prefrontal, anterior cingulate, and default mode regions [40–42], areas that are vulnerable in early AD [43–50]. Loss of awareness may therefore reflect network-level dysfunction or impaired metacognitive processing that is not fully captured by global cerebrospinal fluid biomarker concentrations or clinical decline as shown via our sensitivity analyses. In this context, unawareness may represent a behavioral manifestation of emerging network disintegration, signaling reduced compensatory capacity and increased vulnerability to clinical decline.

From a clinical standpoint, these findings underscore the potential value of incorporating structured informant-based assessments into routine evaluation of MCI. As MCI-stage disease-modifying therapies, such as lecanemab and donanemab, increasingly require accurate prognostication and biomarker confirmation, scalable behavioral markers that help identify individuals at highest risk of progression are critically needed. Awareness phenotyping derived from brief questionnaires may provide complementary information alongside fluid and imaging biomarkers. Future studies are needed to determine its incremental value and clinical utility in diverse populations and clinical settings.

The reduced progression risk observed in the Worried Well phenotype further underscores the clinical relevance of distinguishing directional awareness patterns. Despite comparable biomarker levels to the Low Complaints phenotype, individuals with heightened but uncorroborated concern demonstrated lower progression risk. Heightened awareness may reflect non–Alzheimer-related factors such as anxiety, affective symptoms, or heightened health vigilance rather than underlying neurodegenerative pathology, consistent with prior reports that exaggerated concern is not specific to AD processes [5, 17, 18, 32]. Together, these findings suggest that unawareness may be more closely linked to AD pathology than heightened awareness in MCI.

Several limitations warrant consideration. Although the present study focused on memory self-awareness in individuals with amnestic MCI, future studies should directly compare awareness across memory, executive, language, visuospatial, and other cognitive domains to determine whether the observed associations are specific to memory self-awareness or reflect a broader phenomenon across cognitive domains. Given that validation of the ECog memory subset remains limited outside of ADNI, future work should also establish the clinical validity and generalizability of this measure in independent cohorts. The cohort was predominantly White and highly educated, potentially limiting generalizability. Additionally, only 14% of participants progressed to AD dementia during the follow-up period. Participants who progressed were observed over longer follow-up periods than non-progressors. Additionally, non-random attrition may have influenced the observed progression rates. For example, participants with non-Alzheimer neurodegenerative syndromes, such as frontotemporal dementia, may have been more likely to discontinue follow-up, potentially introducing informative censoring. As with any data-driven clustering approach, replication in independent cohorts will be important to establish the robustness, reproducibility, and generalizability of the identified phenotypes. Although the present study demonstrated consistent associations between memory self-awareness phenotypes, AD biomarkers, and longitudinal clinical progression within ADNI, future work should determine whether these phenotypes can be reproduced in independent cohorts. Regarding the use of ECog, each question asks how the participant’s memory performance compares to 10 years before. The extended recall interval may introduce recall bias if either the participant or the informant inaccurately recalls functioning over the preceding decade, potentially influencing estimates of self-awareness. Additionally, literature has shown that informant scores can be influenced by informant mood, anxiety, and caregiver burden e.g.[51]. That is, if caregiver burden or distress inflates informant-reported complaints, some individuals classified as "Unaware" in our framework may instead reflect elevated caregiver-reported concern driven partly by caregiver factors rather than patient unawareness. Measures of caregiver burden and mood were not available in ADNI and therefore could not be incorporated into the present analyses. Subsequently, it would be important for future studies to incorporate informant-level psychological measures to disambiguate patient unawareness from informant-driven over-reporting. Furthermore, in the unawareness and heightened awareness subscores, individuals with deep complaints (complaints on few questions but high complaints on those questions) can be indistinguishable to individuals with broad complaints (complaints on many questions though low complaint magnitude on each). Future studies should determine whether the breadth and depth of participant–informant discrepancies contribute differentially to disease progression and whether these characteristics should be incorporated into future clustering approaches. Finally, replication in more diverse populations and validation using additional biomarkers, including PET, plasma, and blood-based measures, will be important to establish the robustness, generalizability, and potential clinical utility of these findings.

In conclusion, memory self-awareness phenotypes defined by patient–informant discrepancies were associated with Alzheimer’s disease biomarker burden and differential progression risk in individuals with amnestic MCI. These findings support the biological relevance of memory self-awareness as a marker of clinically meaningful disease heterogeneity while highlighting the need for future studies to determine how awareness across different cognitive domains relates to AD pathology and clinical progression.

## Data Availability

Data for this study are from ADNI and thus is available to the public to download (adni.loni.usc.edu).

## Acknowledgements

Data collection and sharing for this project was funded by the Alzheimer’s Disease Neuroimaging Initiative (ADNI) (National Institutes of Health Grant U01 AG024904) and DOD ADNI (Department of Defense award number W81XWH-12-2-0012). NES had full access to all the data in the study and takes responsibility for the integrity of the data and the accuracy of the data analysis.

## Funding Sources

ADNI is funded by the National Institute on Aging, the National Institute of Biomedical Imaging and Bioengineering, and through generous contributions from the following: AbbVie, Alzheimer’s Association; Alzheimer’s Drug Discovery Foundation; Araclon Biotech; BioClinica, Inc.; Biogen; Bristol-Myers Squibb Company; CereSpir, Inc.; Cogstate; Eisai Inc.; Elan Pharmaceuticals, Inc.; Eli Lilly and Company; EuroImmun; F. Hoffmann-La Roche Ltd and its affiliated company Genentech, Inc.; Fujirebio; GE Healthcare; IXICO Ltd.;Janssen Alzheimer Immunotherapy Research & Development, LLC.; Johnson & Johnson Pharmaceutical Research & Development LLC.; Lumosity; Lundbeck; Merck & Co., Inc.; Meso Scale Diagnostics, LLC.; NeuroRx Research; Neurotrack Technologies; Novartis Pharmaceuticals Corporation; Pfizer Inc.; Piramal Imaging; Servier; Takeda Pharmaceutical Company; and Transition Therapeutics. The Canadian Institutes of Health Research is providing funds to support ADNI clinical sites in Canada. Private sector contributions are facilitated by the Foundation for the National Institutes of Health (www.fnih.org). The grantee organization is the Northern California Institute for Research and Education, and the study is coordinated by the Alzheimer’s Therapeutic Research Institute at the University of Southern California. ADNI data are disseminated by the Laboratory for Neuro Imaging at the University of Southern California. PV was supported by the US National Institute of Aging—NIH grants (R01AG061083, PI: PV).

## Consent Statement

All human subjects provided informed consent.

## Notes

### Competing Interest Statement

N. Enbar-Salo: None to disclose. P. Palmer: None to disclose. C. Munro: Dr. Munro reported receiving a research fellowship grant paid to her institution from the Alzheimer's Association, and serving as Program Chair of the Executive Committee of the Alzheimer's Association ISTAART Neuropsychiatric Syndromes Professional Interest Area, outside the submitted work. M. Dubbelman: None to disclose. A. Aaronson: None to disclose. R. Nosheny: Dr. Nosheny reported receiving grants to her institution from the National Institutes of Health, AbbVie, the Alzheimer's Association, Eli Lilly and Company, Genentech, and the California Department of Public Health; honoraria from the Alzheimer's Therapeutic Research Institute, Washington University in St. Louis, and Duke University; support for meeting travel from the Alzheimer's Therapeutic Research Institute and Washington University in St. Louis; service on the advisory boards of the North Carolina Registry for Brain Health and the International Neurodegenerative Disorders Research Center; chairing the ISTAART Subjective Cognitive Decline Professional Interest Area; and stock options in Vaicro, Inc., all outside the submitted work. J. Gatchel: Dr. Gatchel reported receiving grants from the National Institutes of Health National Institute on Aging during the conduct of the study and, outside the submitted work, consulting fees from Eisai and Oliver Wyman; honoraria from the University of California Irvine Institute for Memory Impairments and Neurological Disorders, the Harvard Radcliffe Institute, and Belvoir; and support for meeting travel from the International Psychogeriatrics Association and the Massachusetts General Hospital Department of Psychiatry Rappaport Fellowship. G. Marshall: Dr Marshall reported receiving grants from Eli Lilly and Company, Eisai, and Genentech outside the submitted work. P. Vannini: Dr. Vannini reported receiving grants from the National Institutes of Health National Institute on Aging during the conduct of the study.

### Author Declarations

Ethics committees or IRBs of the following institutions gave ethical approval for this work, using the Alzheimer's Disease Neuroimaging Initiative (ADNI) dataset: Albany Medical Center Committee on Research Involving Human Subjects Institutional Review Board; Boston University Medical Campus and Boston Medical Center Institutional Review Board; Butler Hospital Institutional Review Board; Cleveland Clinic Institutional Review Board; Columbia University Medical Center Institutional Review Board; Duke University Health System Institutional Review Board; Emory Institutional Review Board; Georgetown University Institutional Review Board; Health Sciences Institutional Review Board; Houston Methodist Institutional Review Board; Howard University Office of Regulatory Research Compliance; Icahn School of Medicine at Mount Sinai Program for the Protection of Human Subjects; Indiana University Institutional Review Board; Institutional Review Board of Baylor College of Medicine; Jewish General Hospital Research Ethics Board; Johns Hopkins Medicine Institutional Review Board; Lifespan - Rhode Island Hospital Institutional Review Board; Mayo Clinic Institutional Review Board; Mount Sinai Medical Center Institutional Review Board; Nathan Kline Institute for Psychiatric Research & Rockland Psychiatric Center Institutional Review Board; New York University Langone Medical Center School of Medicine Institutional Review Board; Northwestern University Institutional Review Board; Oregon Health and Science University Institutional Review Board; Partners Human Research Committee; Research Ethics Board, Sunnybrook Health Sciences Centre; Roper St. Francis Healthcare Institutional Review Board; Rush University Medical Center Institutional Review Board; St. Joseph's Phoenix Institutional Review Board; Stanford Institutional Review Board; The Ohio State University Institutional Review Board; University Hospitals Cleveland Medical Center Institutional Review Board; University of Alabama Office of the IRB; University of British Columbia Research Ethics Board; University of California Davis Institutional Review Board Administration; University of California Los Angeles Office of the Human Research Protection Program; University of California San Diego Human Research Protections Program; University of California San Francisco Human Research Protection Program; University of Iowa Institutional Review Board; University of Kansas Medical Center Human Subjects Committee; University of Kentucky Medical Institutional Review Board; University of Michigan Medical School Institutional Review Board; University of Pennsylvania Institutional Review Board; University of Pittsburgh Institutional Review Board; University of Rochester Research Subjects Review Board; University of South Florida Institutional Review Board; University of Southern California Institutional Review Board; University of Texas Southwestern Medical Center Institutional Review Board; Veterans Affairs Long Beach Healthcare System Institutional Review Board; Vanderbilt University Medical Center Institutional Review Board; Wake Forest School of Medicine Institutional Review Board; Washington University School of Medicine Institutional Review Board; Western Institutional Review Board; Western University Health Sciences Research Ethics Board; and Yale University Institutional Review Board. This work used de-identified archived data from ADNI, accessed through a data access request submitted to ADNI (adni.loni.usc.edu) and approved electronically. The ADNI study team obtained all IRB approvals and met all ethical standards in the collection of the data, and written informed consent was obtained from all participants.

